# Multimodal AI fuses proteomic and EHR data for rational prioritization of protein biomarkers in diabetic retinopathy

**DOI:** 10.64898/2026.02.23.26346903

**Authors:** Jonathan B. Lin, Samson J. Mataraso, Madhumeeta Chadha, Gabriel Velez, Prithvi Mruthyunjaya, Nima Aghaeepour, Vinit B. Mahajan

## Abstract

**Purpose:** There is a need for novel therapies for diabetic retinopathy (DR) because existing therapies treat only certain features of DR and do not work optimally for all patients. While proteomic studies provide insight into disease pathobiology, they are often limited to small sample sizes due to high costs, limiting their generalizability and reproducibility. Moreover, they often yield lists of tens to hundreds of proteins with differential expression, making it difficult to prioritize the most biologically relevant biomarkers beyond using arbitrary fold-change and false-detection rate cutoffs. Here, we applied a two-stage multimodal AI approach: first, we integrated EHR and proteomics data to rationally prioritize candidate protein biomarkers and, next, validated these biomarkers in an independent cohort. These protein biomarkers of DR are rooted in the EHR data and thereby more likely to be biological drivers of disease.

**Methods:** We obtained EHR data from a large number of patients with and without DR (N=319,997) from the STARR-OMOP database and obtained aqueous humor liquid biopsies from a subset of these patients (N=101) for high-resolution proteomic profiling. We developed **C**linical and **O**mics **M**ulti-Modal Analysis **E**nhanced with **T**ransfer Learning (COMET) to perform integrated analysis of proteomics and all available EHR data to identify protein biomarkers of DR. The model was trained in two phases: first, it was pretrained using patients with EHR data alone (N=319,896), and then, it was fine tuned using patients with both EHR and proteomics data (N=101), allowing it to learn both clinical and molecular features associated with DR. Findings from COMET were then validated with liquid biopsies from an independent, validation cohort (N=164).

**Results:** t-distributed stochastic neighbor embedding (t-SNE) analysis of EHR and proteomics data identified proteins clustering with related EHR features. Levels of STX3 and NOTCH2, proteins involved in retinal function, were correlated with a diagnosis of macular edema, a record of a visual field exam, and a prescription for latanoprost, highlighting protein-EHR alignment. The pretrained, multimodal COMET model was superior (AUROC=0.98, AUPRC=0.91) compared to models generated using either EHR or proteomics data alone or without pretraining (AUROC: 0.76 to 0.92; AUPRC: 0.47 to 0.74). The proteins SERPINE1, QPCT, AKR1C2, IL2RB, and SRSF6 were prioritized by the COMET model compared to the models without pretraining, supporting their potential role in DR pathobiology, and were subsequently validated in an independent cohort.

**Conclusion:** We used multimodal AI to prioritize protein biomarkers of DR that are most strongly linked to EHR elements, as well as identifying other protein biomarkers associated with disease features like diabetic macular edema. These findings serve as a foundation for future mechanistic studies and highlight the synergistic value of using multimodal AI to fuse EHR and proteomics data for enhanced proteomics analysis.

## INTRODUCTION

Diabetic retinopathy (DR) is a leading cause of blindness worldwide. While numerous systemic treatments for diabetes, including the newest glucagon-like peptide-1 (GLP1) receptor agonists, have improved glycemic control in patients and, thereby, have reduced the burden of diabetes-related microvascular retinal damage and associated vision loss ^1–4^, recent population studies demonstrate that visual morbidity from DR remains prevalent, especially among racial and ethnic minority groups ^5,6^. For patients who develop features of advanced disease, such as diabetic macular edema (DME) or proliferative DR (PDR), intraocular agents that target vascular endothelial growth factor (VEGF), intraocular corticosteroids, laser photocoagulation, or sometimes, vitreoretinal surgery, can slow or halt disease progression ^1^. However, a significant proportion of patients demonstrate limited response or fail to respond to these existing treatments, highlighting that there are heterogeneous biological pathways driving disease pathobiology that go beyond VEGF signaling ^7,8^. Therefore, it is crucial to deepen our understanding of the mechanisms driving DR to develop VEGF-independent therapies. These treatments could be used for patients who do not respond to current therapeutic options and may even allow for earlier intervention to prevent DME and PDR.

Omics has become a widely adopted approach for uncovering the cellular and molecular mechanisms contributing to disease. These studies interrogate which genes, gene transcripts, proteins, metabolites, or molecules are altered in disease states. Highly sensitive, advanced assays can characterize the proteome from as little as a few hundred microliters of liquid ^9^.

Nonetheless, interpreting the large amount of data generated by omics studies is a significant challenge since it requires sifting through large numbers of analytes to identify clinically relevant molecular markers and drivers of disease ^10^. Manually selecting which genes or molecules to focus on is often influenced by an investigator’s own hypotheses or prior work, potentially overlooking the most clinically relevant analytes. Moreover, due to high cost, omics studies are usually done with sample sizes ranging from tens to hundreds, which can limit their generalizability and reproducibility ^11,12^.

The digitization of modern medicine has generated a wealth of electronic health record (EHR) data that may be leveraged to enhance our understanding of human disease ^13^. For instance, the American Academy of Ophthalmology Intelligent Research in Sight (IRIS) Registry, the largest such patient database in the world, has provided insights into longitudinal trends in treatment patterns for DR ^14^. Advantages of EHR-based studies are they do not require study recruitment, they include elements of clinical examination and treatment outcomes, something not found in coding-based databases (e.g, the Medicare 5% Sample, Truven, TriNetX), and they capture extremely large patient populations, often numbering in the thousands to millions.

Nonetheless, studies based solely on EHR data lack the depth to identify molecular or cellular drivers of disease, which is necessary for developing novel therapeutics.

Previous proteomics studies have contributed greatly to our understanding of the vast molecular changes exhibited in DR, for example, by identifying proteins that may be linked to progression to proliferative DR ^15^. However, it remains difficult to prioritize the individual proteins that are drivers of disease among the tens to hundreds of proteins that may be differentially expressed, leading to reliance on arbitrary fold-change and false detection rate cutoffs. An unmet need is to use multimodal AI to incorporate EHR data to allow for more rational protein prioritization. In other words, fusing EHR and molecular omics data may enable more sophisticated selection of proteins in an unbiased manner. Previous approaches for integrating EHR and omics data were often limited by the need for complete data across all modalities or by challenges in learning cross-modal interactions ^16^. We recently developed a new integrative methodology termed **C**linical and **O**mics **M**ulti-Modal Analysis **E**nhanced with **T**ransfer Learning (COMET), which overcomes these limitations ^17^. COMET is a multimodal, deep learning architecture that leverages EHR data from large populations for pretraining to enhance omics analysis.

In this study, we applied COMET to fuse EHR and proteomics data from a large cohort of patients with DR with the goal of rationally prioritizing proteins that may be biologically most relevant in DR among the hundreds of proteins that have differential expression in DR and subsequently validated these proteins with an independent cohort. Our multimodal AI approach identified key proteins that are associated with specific disease features, such as macular edema, and other key proteins perturbed in DR that are most strongly linked to EHR data, some of which may have been overlooked by conventional biomarker discovery approaches.

Cumulatively, our findings not only serve as a foundation for future mechanistic studies but also highlight the synergistic value of using a multimodal AI-based approach to analyze EHR and proteomics data.

## RESULTS

We identified a large cohort of 319,997 patients, both with and without DR, who received their clinical care at a single academic medical center. We extracted all of the available structured EHR data for these patients from the **STA**nford **R**esearch data **R**epository (STARR) **O**bservational **M**edical **O**utcomes **P**artnership (OMOP) database. The OMOP format is a standardized data model that contains EHR data in tabular form and includes information about measurements, observations, drug exposures, condition occurrences, and procedure occurrences in a dichotomous manner (i.e., present versus absent). For each patient, there were 2,531 EHR features for a total of over 809 million EHR data points. We embedded longitudinal EHR data using word2vec, as described previously ^17^. The complete list of EHR features is available in **Supplementary Data 1**. This approach of analyzing all of the available EHR data rather than selectively choosing a small number of clinical features allowed us to perform hypothesis-agnostic analysis and to minimize any potential bias. For a subset of these patients (N=101; 14 with DR [5 non-proliferative and 9 proliferative] and 87 without DR; “discovery cohort”), we collected aqueous humor liquid biopsies at the time of planned ocular surgery. Demographic and clinical characteristics of all these patients are summarized in **Table 1**. We then performed high-resolution proteomic profiling of the liquid biopsies using an aptamer-based platform, as described previously ^18^.

**Table 1.**
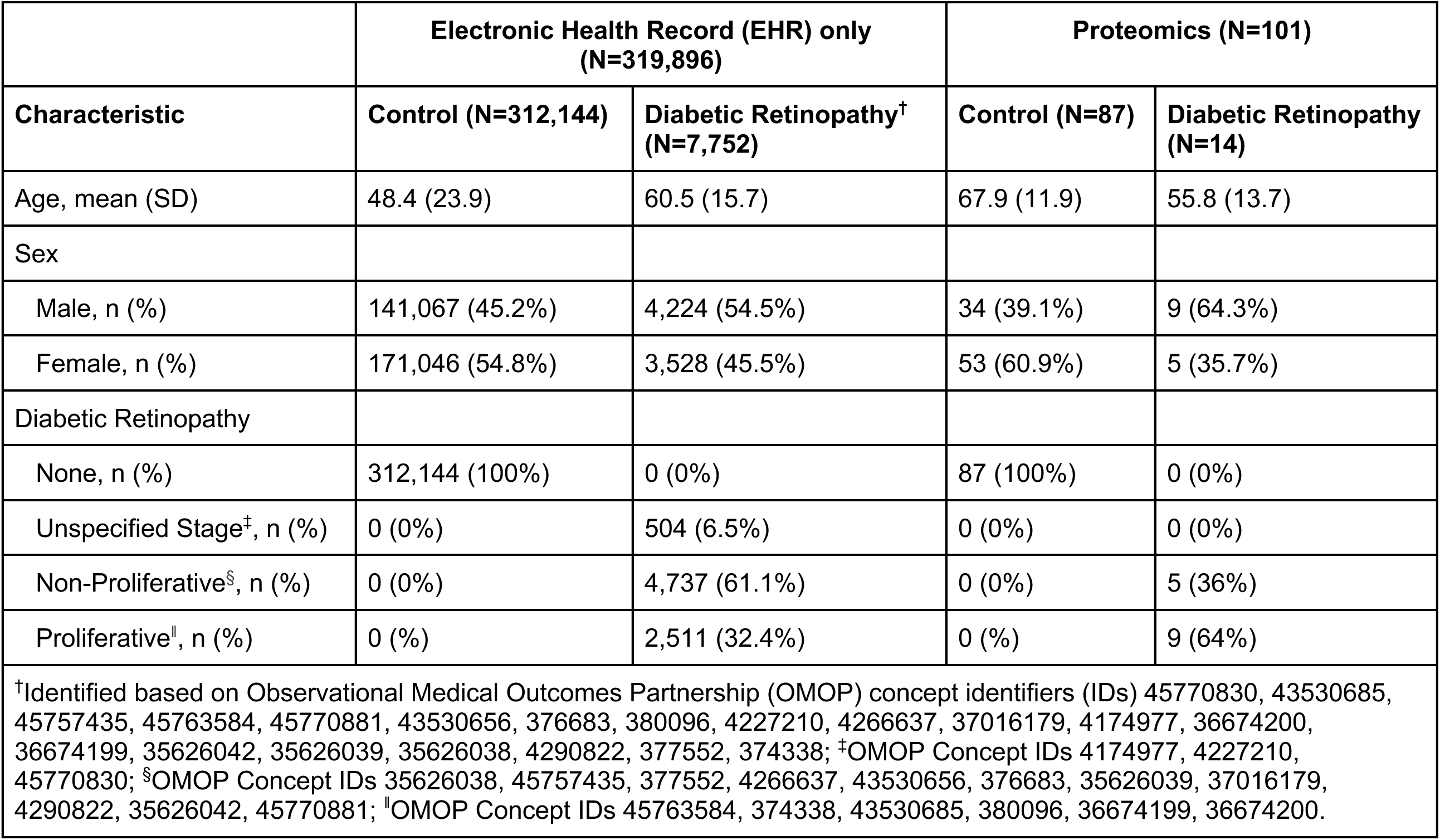
Demographic and clinical information of participants in the discovery cohort.

### There is alignment between the electronic health record (EHR) and proteomics data

To determine whether there was alignment between the EHR and proteomics data, we performed a t-distributed stochastic neighbor embedding (t-SNE) analysis using the embedded longitudinal EHR data and the proteomics data from patients with both data types (**Figure 1a**). We used a correlation matrix of all variables for t-SNE analysis to help scale the data. In this visualization, each circle represents an individual EHR feature or proteomics variable. Circles located in close proximity to each other have similar correlations to each other and other nearby variables. A complete list of the variables shown in **Figure 1a** is provided in **Supplementary Data 1**.

**Figure 1.**
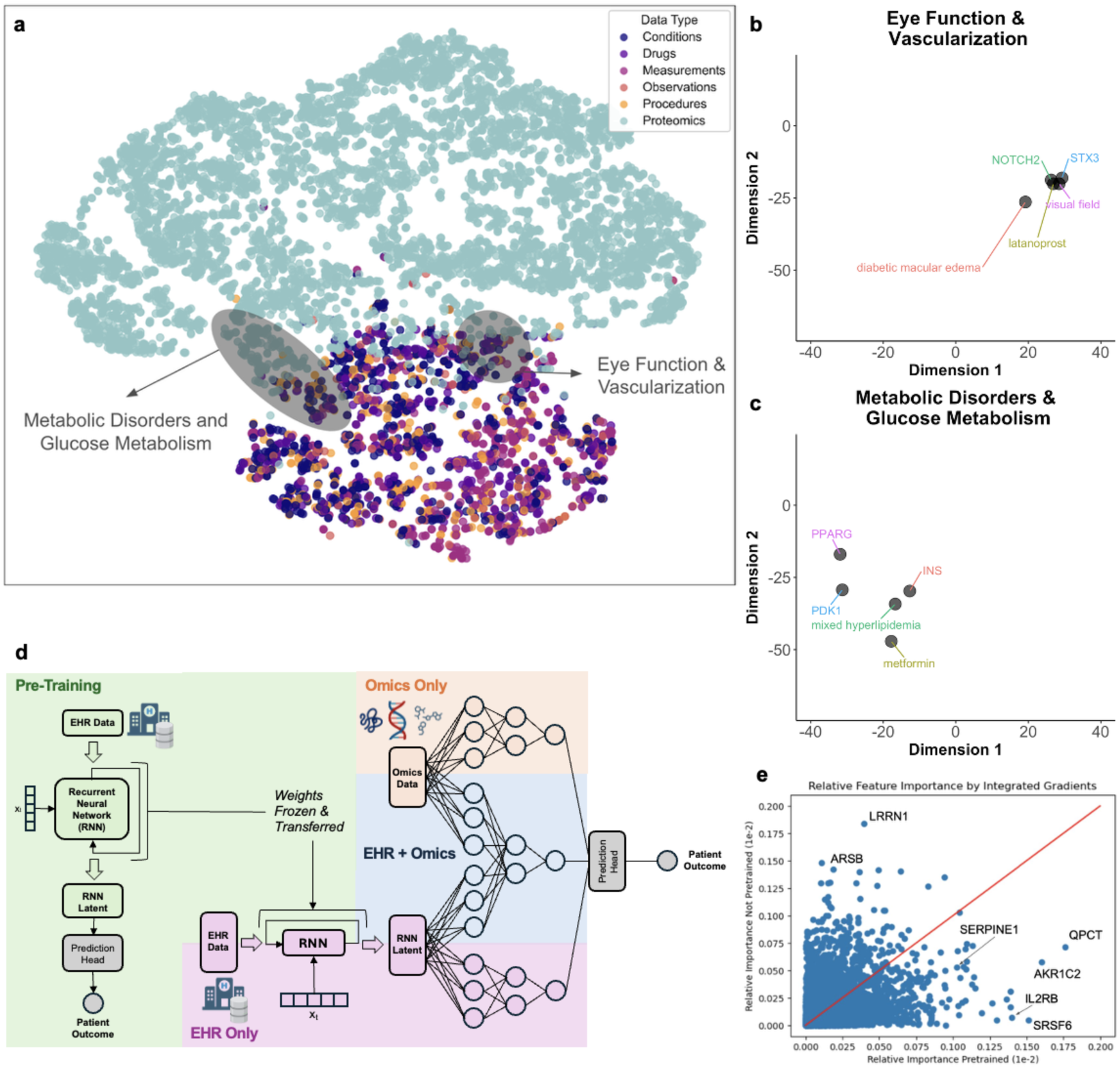
(a) Visualization of multimodal data using t-distributed stochastic neighbor embedding (t-SNE). The correlation matrix is projected into two unitless dimensions, and each circle represents an individual electronic health record (EHR) or proteomics feature. Circles that cluster in the same physical space have similar correlations with other surrounding variables. (b) One cluster we labeled “*Eye Function & Vascularization*” includes proteins syntaxin 3 (STX3) and neurogenic locus notch homolog protein 2 (NOTCH2), along with EHR features related to the retina and ocular function. (c) Another cluster we labeled “*Metabolic Disorders & Glucose Metabolism*” includes proteins insulin (INS), pyruvate dehydrogenase kinase 1 (PDK1), and peroxisome proliferator-activated receptor gamma (PPARG), and EHR features related to metabolic disease. (d) Schematic of the **C**linical and **O**mics **M**ulti-Modal Analysis **E**nhanced with **T**ransfer Learning (COMET) model architecture. The electronic health record (EHR) data is used in a pretraining stage with the core architecture of a recurrent neural network (RNN) (green shading). After pretraining, the RNN weights are frozen and transferred into a multimodal architecture that analyzes both EHR and omics data. The orange shading depicts a conventional omics-only pipeline; the purple shading depicts a conventional EHR-only pipeline; and the blue shading depicts a combined EHR and omics pipeline without pretraining. (e) Some proteins were prioritized in the COMET model versus the non-pretrained model. Each blue circle represents an individual protein. Blue circles in the lower right, including SERPINE1, QPCT, AKR1C2, IL2RB, and SRSF6, had higher relative importance in the pretrained models.

Through visual inspection, we identified clusters of proteins previously reported to play an important role in the ocular physiology, along with related EHR features. One such cluster, which we labeled “*Eye Function and Vascularization,*” included the proteins syntaxin 3 (STX3) and neurogenic locus notch homolog protein 2 (NOTCH2), as well as the diagnosis of diabetic macular edema, a record of a visual field exam, and a prescription for latanoprost (**Figure 1b**).

Both STX3 and NOTCH2 have established roles in retinal function and vision. In murine models, deletion of STX3 disrupts the trafficking of photoreceptor outer segment membrane proteins ^19^, and in humans, STX3 mutations are associated with retinal neurodegeneration ^20^. Notably, NOTCH2 not only has a role in retinal development ^21^, but it can also induce vascular permeability ^22^, a mechanism by which it may contribute to the pathogenesis of diabetic macular edema.

Another cluster, which we labeled “*Metabolic Disorders and Glucose Metabolism,*” included the proteins insulin (INS), pyruvate dehydrogenase kinase 1 (PDK1), and peroxisome proliferator-activated receptor gamma (PPARG), along with a diagnosis of mixed hyperlipidemia and a prescription for metformin (**Figure 1c**). INS, PDK1, and PPARG are well established regulators of systemic metabolic regulation ^23^. Collectively, these findings underscore the strong concordance between the proteomics and EHR-derived clinical features.

### Pretraining proteomics analysis with EHR data enhances model performance

Next, we examined whether fusing proteomics data with all available EHR data could enhance biological discovery and enable prioritization of more biologically relevant proteins. COMET is a deep learning framework designed to improve the interpretation of omics studies by leveraging EHR data from large patient populations ^17^. In brief, COMET first pretrains a model using structured EHR data from a large, observational cohort without omics data and then transfers this knowledge to a multimodal architecture applied to patients with both EHR and omics data, thereby optimizing model performance. A schematic comparing the COMET architecture to conventional pipelines, including EHR-only, proteomics-only, and EHR plus proteomics without transfer learning, is shown in **Figure 1d**.

We developed four separate models using 1) EHR data alone (N=101), 2) proteomics data alone (N=101), 3) combined EHR and proteomics data without pretraining (N=101), and 4) combined EHR and proteomics data with pretraining using COMET (N=319,896 for pretraining, N=101 for fine tuning and evaluation). A rigorous bootstrapped, Monte Carlo cross-validation approach with twenty-five independent iterations was used to ensure that the findings were robust. The average performance metrics from twenty-five iterations of these models are shown in **Table 2**. The Area Under the Receiver Operating Characteristic Curve (AUROC), a measure of model discrimination, was highest for COMET (AUROC: 0.98; 95% confidence interval [CI]: 0.92 to 1.00), outperforming the EHR-alone (AUROC: 0.76; 95% CI: 0.59 to 0.90), the proteomics-alone (AUROC: 0.92; 95% CI: 0.83 to 0.99), and the EHR plus proteomics without pretraining models (AUROC: 0.92; 95% CI: 0.84 to 0.98). Similarly, COMET achieved superior performance based on Area Under the Precision-Recall Curve (AUPRC), another measure of model discrimination (AUPRC: 0.91; 95% CI: 0.77 to 1.00) when compared to the EHR-alone (AUPRC: 0.47; 95% CI: 0.24 to 0.74), the proteomics-alone (AUPRC: 0.74; 95% CI: 0.49 to 0.94), and the combined, not pretrained models (AUPRC: 0.73; 95% CI: 0.49 to 0.91). These results demonstrate that COMET’s ability to fuse EHR and proteomics data with pretraining significantly enhances model performance and may offer a powerful approach for advancing biological discovery.

**Table 2.** Performance metrics for models of proteins perturbed in diabetic retinopathy using different combinati health record (EHR) and proteomics data with and without pretraining. AUROC: **A**rea **U**nder the **R**eceiver **O**per curve; AUPRC = **A**rea **U**nder the **P**recision **R**ecall **C**urve; COMET: **C**linical and **O**mics **M**ulti-Modal Analysis **E**n Learning; 95% CI: 95% confidence interval (calculated from one thousand iterations of bootstrapping).

| Model | Features | Sample Size | AUROC<br>[95% CI] | AUPRC<br>[95% CI] |
| --- | --- | --- | --- | --- |
| 1 | EHR only | 101 | 0.76<br>[0.59,0.90] | 0.47<br>[0.24, 0.74] |
| 2 | Proteomics only | 101 | 0.92<br>[0.83, 0.99] | 0.74<br>[0.49, 0.94] |
| 3 | EHR and proteomics, no pretraining | 101 | 0.92<br>[0.84, 0.98] | 0.73<br>[0.49, 0.91] |
| 4 | COMET (EHR and proteomics with pretraining) | 319,896 for pretraining, 101 for fine tuning and evaluation | 0.98<br>[0.92, 1.00] | 0.91<br>[0.77, 1.00] |

### The pretrained EHR-proteomics model prioritizes biologically and clinically relevant proteins in diabetic retinopathy

Because the COMET-trained model achieved the highest performance metrics, we conducted a post hoc analysis to identify the proteins prioritized by the pretrained COMET EHR-proteomics model in comparison with the EHR-proteomics model without pretraining. We evaluated feature importance using the integrated gradients method ^24^. We were particularly interested in proteins that had higher feature importance in the COMET model in comparison with the baseline proteomics-alone model, as these are more rooted in the EHR data and, therefore, may be central in disease pathobiology. To this end, we compared the relative feature importance of all proteins in the pretrained COMET model versus the baseline, non-pretrained model (**Figure 1e**). The proteins SERPINE1, QPCT, AKR1C2, IL2RB, and SRSF6 exhibited the largest fold change in feature importance (**Table 3**). While several of these proteins have previously been associated with DR, they have not been prioritized nor have they been the focus of novel targeted therapeutics to our knowledge. Notably, because DR stage (mild, moderate, or severe nonproliferative versus proliferative) and glycemic control (hyperglycemia) are EHR features that are captured within STARR OMOP, the protein biomarkers highlighted in this integrative analysis are not specific to a single DR stage and are independent of glycemic control.

**Table 3.**
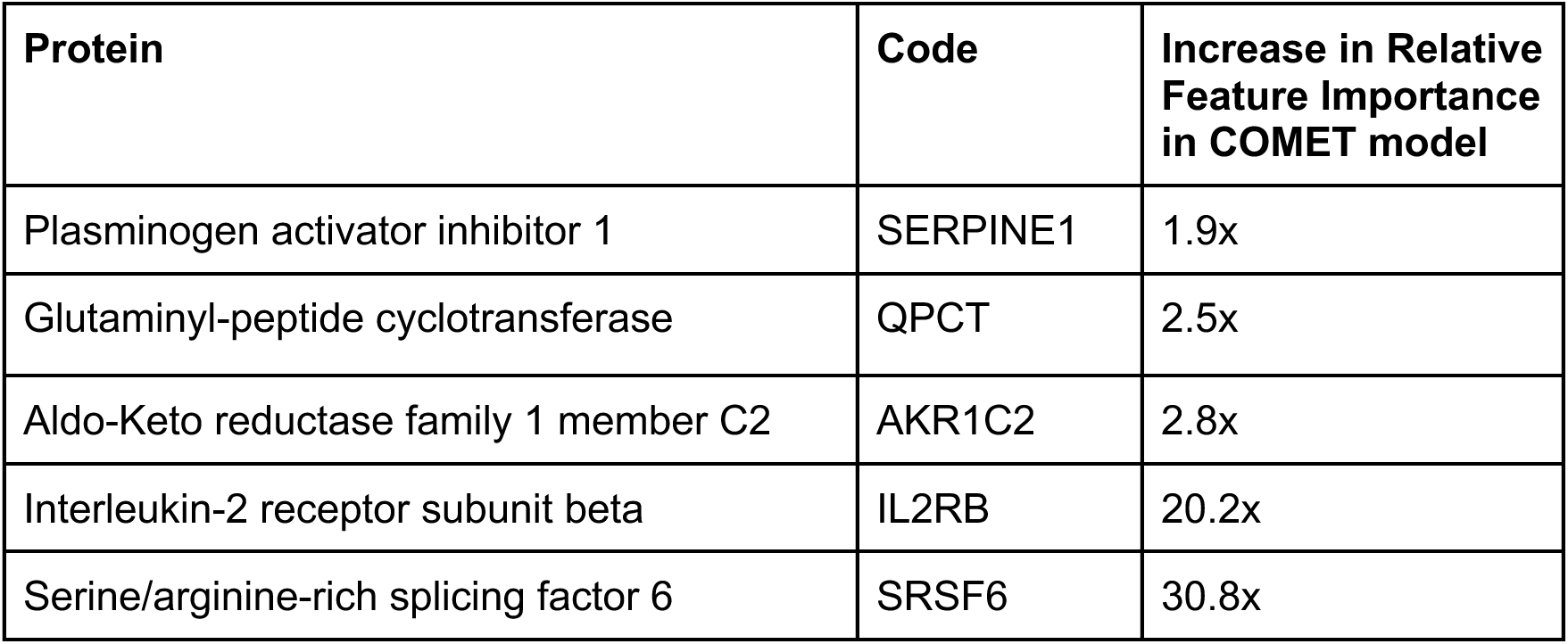
Proteins identified as more important in pretrained COMET model for diabetic retinopathy.

SERPINE1 (plasminogen activator inhibitor 1; also known as PAI1) inhibits plasminogen activator and is, thus, a prothrombotic agent that has been associated with obesity, insulin resistance, and increased vascular risk. Glutaminyl-peptide cyclotransferase (QPCT) is an enzyme that plays a key role in the posttranslational modifications of neuroendocrine peptides. Aldo-keto reductase family 1 member C2 (AKR1C2) is an enzyme that catalyzes the conversion of aldehydes and ketones to their corresponding alcohols. Interleukin 2 receptor subunit beta (IL2RB) is a component of the IL2 receptor complex, which plays a central role in regulating T cell function within the adaptive immune system. Finally, serine/arginine-rich splicing factor 6 (SFSF6) is involved in regulation of alternative splicing of precursor RNA molecules.

Notably, two of these five proteins, AKR1C2 and IL2RB, could have been missed by applying standard differential expression analysis parameters, e.g., false detection rate (FDR) < 0.05 and | log_2_(fold change) | >2), as a strategy for prioritizing proteins instead of using COMET. Power calculations reveal that to achieve 90% statistical power to detect the magnitude of difference observed in these two proteins, it would have required much larger sample sizes of N=143/group and N=45/group, respectively. As such, applying multimodal AI to incorporate EHR data into proteomics analysis can prioritize biologically relevant protein biomarkers and enable identification of important proteins with smaller sample sizes.

As previously discussed, a key challenge of small-scale omics studies is poor reproducibility. To determine whether COMET could improve the reproducibility of proteomics findings, we sought to validate our proteomic findings in an independent cohort. To achieve 90% statistical power to detect significant differences similar in magnitude to those identified in our discovery cohort (d=1.4), we calculated the need for 127 controls and 7 patients with DR. To ensure rigor, the validation cohort exceeded this sample size and consisted of N=164 participants (17 with DR and 147 without). As above, we collected aqueous humor liquid biopsies from these patients at the time of planned ocular surgery. Proteomics profiling of these samples was performed using the same high-resolution, aptamer-based platform as in the discovery cohort. Demographic and clinical characteristics of the validation cohort are summarized in **Supplementary Data 2**. We examined the levels of the five proteins with the highest relative feature importance, SERPINE1, QPCT, AKR1C2, IL2RB, and SRSF6, and found similar patterns of statistically significant differences in DR versus control across both discovery and validation cohorts (**Figures 2a-b**).

**Figure 2.**
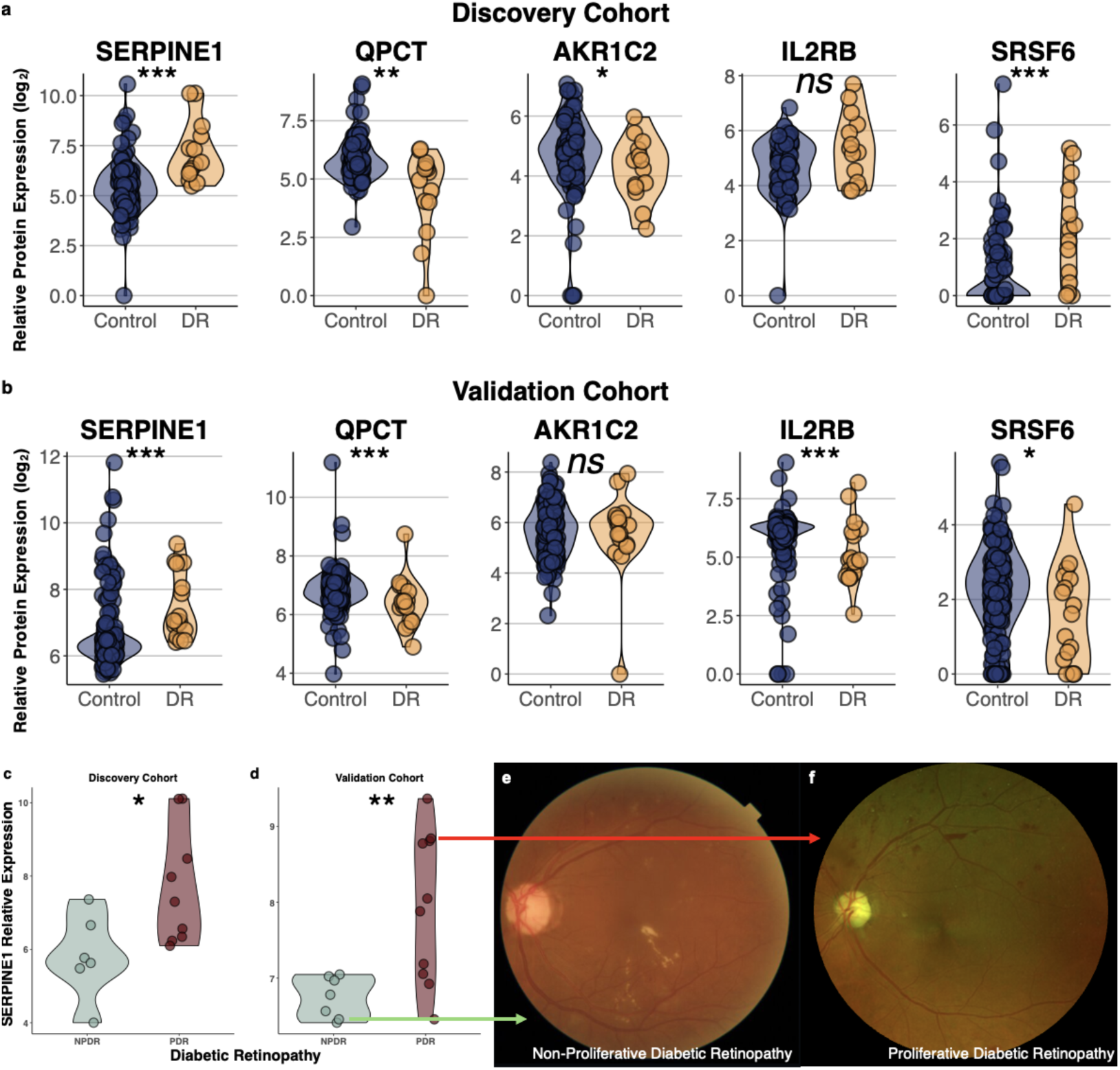
(a-b) Violin plots showing relative aqueous humor levels of the proteins that had the highest relative feature importance in the COMET model compared to the non-pretrained model. Similar patterns were seen when comparing patients with diabetic retinopathy (DR) (yellow) to control patients without DR (blue) in both the (a) discovery cohort (N=101; N=14 DR, N=87 control) and the (b) validation cohort (N=164; N=17 DR, N=147 control). (c-d) Relative aqueous humor SERPINE1 levels (log_2_-transformed) were significantly higher in patients with proliferative DR (PDR) compared to non-proliferative DR (NPDR) in both the (c) discovery cohort and the (d) validation cohort. (e) Representative fundus photograph of patient from the validation cohort with the second lowest SERPINE1 level demonstrates scattered exudates, particularly along the inferior arcade, and rare dot-blot hemorrhages along the superior arcade, consistent with NPDR, and retinal thickening consistent with diabetic macular edema. (f) Representative fundus photograph of patient from the validation cohort with the third highest SERPINE1 level demonstrates numerous dot-blot hemorrhages, intraretinal microvascular abnormalities, and foci of neovascularization elsewhere (NVE), consistent with PDR. This patient subsequently developed non-clearing vitreous hemorrhage that required pars plana vitrectomy. Individual circles depict individual patients (a-d). * p < 0.05; ** p < 0.01; *** p < 0.001; ns: not significant (a-d).

As a proof of concept, we also sought to determine whether any of the proteins with highest relative feature importance in the COMET model may be associated with certain features of disease, as this may serve as the basis for novel therapeutics. We substratified the patients with DR into those with NPDR versus PDR through masked retrospective review of the medical record. In both the discovery cohort, as well as the validation cohort, SERPINE1 levels were significantly higher in patients with PDR compared to those with NPDR (**Figures 2c-d**). The patient with the second lowest SERPINE1 level from the validation cohort had moderate NPDR and was receiving intravitreal bevacizumab pro re nata for diabetic macular edema (**Figure 2e**). In contrast, the patient with the third highest SERPINE1 levels from the validation cohort had PDR: he had experienced a vitreous hemorrhage in the setting of neovascularization five months prior to aqueous humor collection and subsequently required pars plana vitrectomy and endolaser panretinal photocoagulation (**Figure 2f**). Though classifying patients as NPDR versus PDR is not a diagnostic challenge, the ability to identify the proteins that differ by DR stage may enhance our understanding of the molecular pathways contributing to disease, thereby highlighting the value of fusing proteomic and EHR data for biological discovery.

### The pretrained EHR-proteomics model yields an EHR latent representation rooted in biology

Next, we investigated whether incorporating proteomics data within the COMET framework improves the modeling of the EHR data since one criticism of EHR data is that it is not rooted in a biological basis. To do this, we compared the alignment between the proteomics and EHR data using t-SNE analysis. Specifically, we overlaid a t-SNE projection of the raw proteomics data with the 400-dimension latent representation of the EHR data generated with model #3 (EHR and proteomics data without pretraining) (**Figure 3a**) and compared it to the same projection generated by model #4 (COMET) (**Figure 3b**). Lines connecting proteins to EHR features represent statistically significant correlations (**Figures 3a-3b**). Notably, the average absolute correlation between the EHR latent dimensions and proteomic features was significantly higher in the COMET model (mean R = 0.21) compared to the baseline model (mean R = 0.08) (p < 0.0001; **Figure 3c**). These results suggest that COMET enables more meaningful integration of proteomics and EHR data, potentially capturing latent structure in the EHR data that reflects underlying molecular mechanisms.

**Figure 3.**
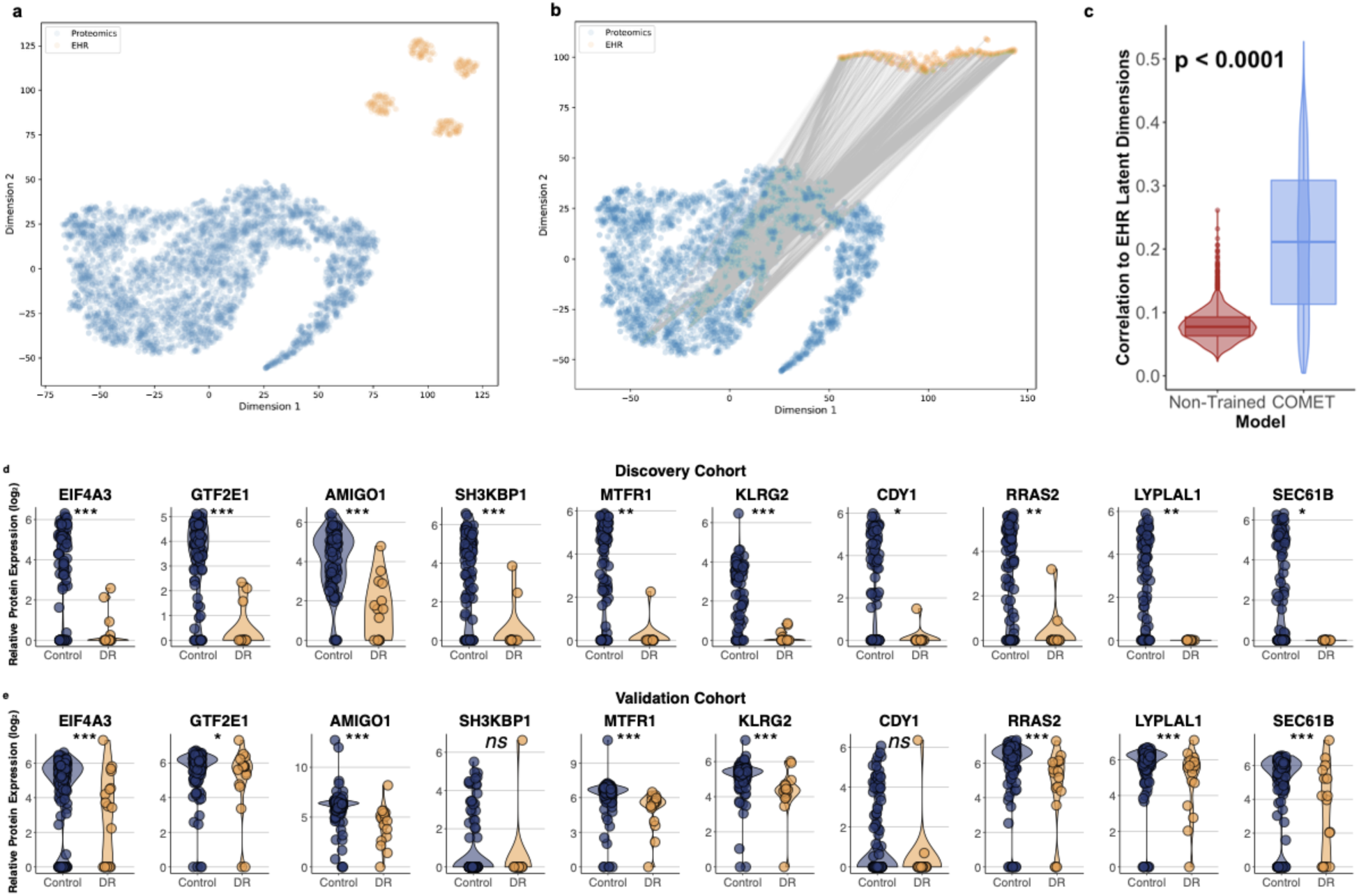
(a-b) t-distributed stochastic neighbor embedding (t-SNE) visualization of the proteomics data (blue circles) and electronic health record (EHR) latent representations (orange circles) from (a) the model generated using the EHR and proteomics data without pretraining compared to (b) that generated using COMET. Gray lines connect statistically significant correlations between individual proteins and EHR latent representation dimensions. The non-pretrained model (a) had zero significant correlations while the COMET model (b) had 4,564 significant correlations, highlighting increased alignment of the EHR data with proteomics data by using COMET. (c) There was significantly higher average absolute correlation between each protein and the EHR latent representations with COMET compared with the non-trained model. (d-e) Violin plots showing relative aqueous humor levels of the proteins that were most significantly correlated with electronic health record (EHR) latent representations in the COMET model. Similar patterns were seen when comparing patients with diabetic retinopathy (DR) (yellow) to control patients without DR (blue) in both the (d) discovery cohort (N=101; N=14 DR, N=87 control) and the (e) validation cohort (N=164; N=17 DR, N=147 control). Individual circles depict individual patients (d-e). * p < 0.05; ** p < 0.01; *** p < 0.001; ns: not significant (d-e).

The proteins most strongly correlated with the EHR latent representations in the COMET model are listed in **Table 4**. Similar patterns of statistically significant differences in DR versus control were seen in these proteins across both the discovery and validation cohorts (**Figures 3d-e**).

**Table 4.** Proteins most significantly correlated with electronic health record (EHR) latent representations in pret for diabetic retinopathy.

| Protein | Code | Percentage of the 400 EHR latent dimensions that were correlated with the protein |
| --- | --- | --- |
| Eukaryotic initiation factor 4A-III | EIF4A3 | 78.50% |
| General transcription factor IIE subunit 1 | GTF2E1 | 77.75% |
| Adhesion molecule with Ig like domain 1 | AMIGO1 | 76.50% |
| SH3 domain-containing kinase-binding protein 1 | SH3KBP1 | 73.75% |
| Mitochondrial fission regulator 1 | MTFR1 | 73.50% |
| Killer Cell Lectin Like Receptor G2 | KLRG2 | 72.00% |
| Chromodomain Y-Linked 1 | CDY1 | 71.75% |
| Ras-related protein R-Ras2 | RRAS2 | 71.00% |
| Lysophospholipase like 1 | LYPLAL1 | 70.75% |
| SEC61 translocon subunit beta | SEC61B | 69.25% |

These proteins participate in fundamental cellular processes, suggesting that COMET captures biologically meaningful patterns within EHR data. For example, eukaryotic initiation factor 4A-III (EIG4A3) and general transcription factor IIE subunit 1 (GTF2E1) are involved in RNA processing and transcription ^25,26^. Adhesion molecule with Ig like domain 1 (AMIGO1) plays a role in cell adhesion and neural development ^27^. SH3 domain-containing kinase-binding protein 1 (SH3KBP1), killer cell lectin like receptor G2 (KLRG2), and ras-related protein R-Ras2 are central in cell signaling and have been implicated in oncogenic process ^28–30^. SEC61 translocon subunit beta (SEC61B) forms a part of the protein translocation channel in the endoplasmic reticulum ^31^. Mitochondrial fission regulator 1 (MTFR1) and lysophospholipase like 1 (LYPLAL1) are involved in mitochondrial function and lipid metabolism, respectively ^32,33^. Finally, chromodomain Y-Linked 1 (CDY1) is a testis-specific protein involved in spermatogenesis ^34^.

Finally, we sought to determine the cellular origin of the proteins highlighted by COMET using another technique we previously developed called TEMPO (**T**racing **E**xpression of **M**ultiple **P**rotein **O**rigins) ^18^. In brief, TEMPO combines liquid-biopsy proteomics with single-cell transcriptomics using AI to trace proteins detected in aqueous humor back to their cellular origin within the eye. We identified fourteen proteins highlighted by COMET that could be traced back to specific ocular cell types, including retinal neurons, immune cells, and vascular cells (**Figure 4**). These findings not only highlight the multifactorial contributions of retinal neuroinflammation, microvascular damage, and neurodegeneration in DR pathobiology but also indicate there are cell-autonomous mechanisms within the eye contributing to DR pathophysiology beyond the systemic burden of disease.

**Figure 4.**
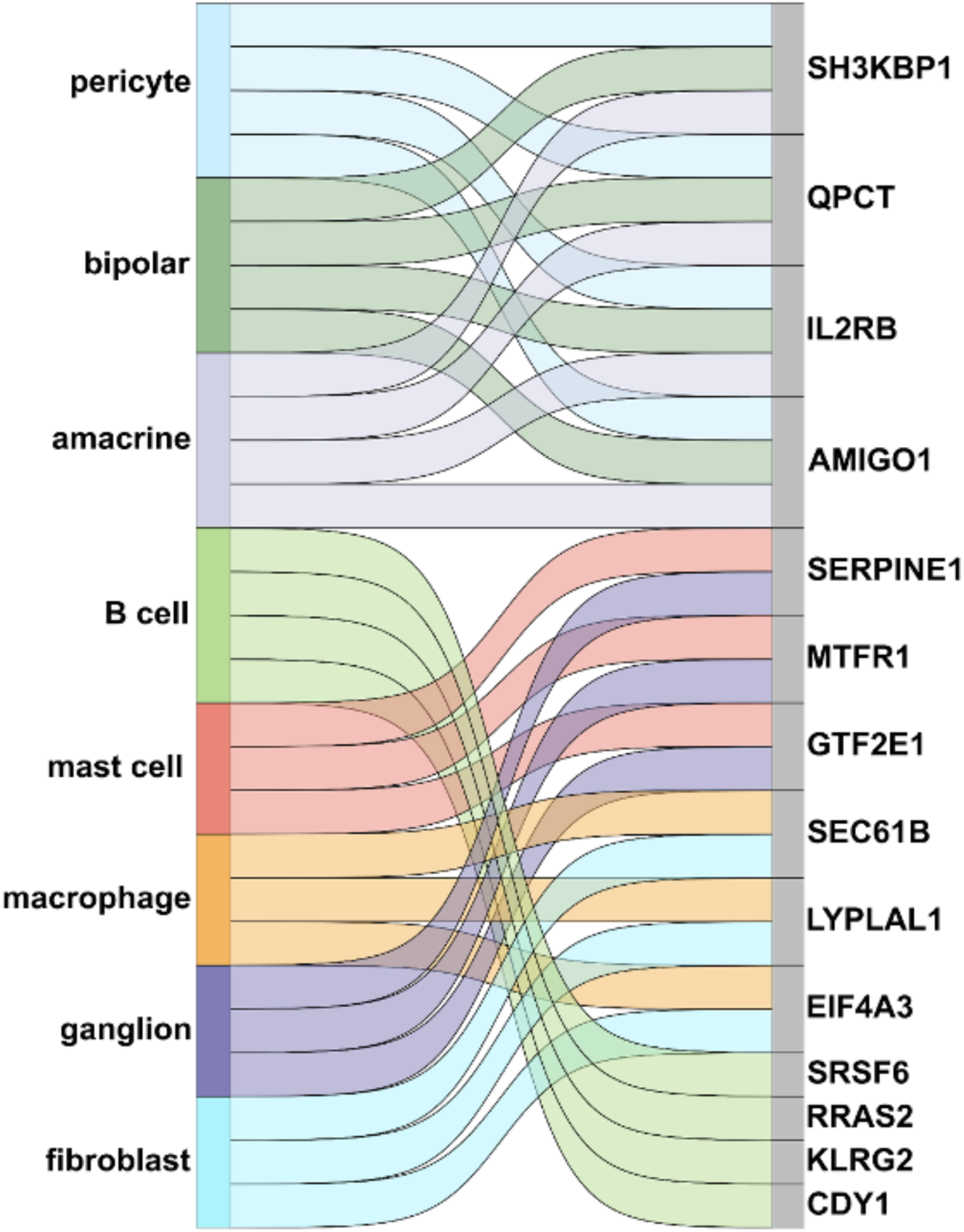
Sankey plot depicts the diverse cellular origins of the protein biomarkers of diabetic retinopathy (DR) prioritized by multimodal AI approach to integrate electronic health record (EHR) data, suggesting possible cell-autonomous mechanisms within the eye contributing to DR pathophysiology beyond the systemic burden of disease. A cell type was considered to be the origin of the protein if the corresponding gene expression was at least two standard deviations above the mean expression in all cell types.

## DISCUSSION

Here, we applied COMET, an innovative, deep-learning approach, which uses transfer learning to integrate large retrospective EHR databases with proteomics data, to prioritize the most clinically relevant protein biomarkers of DR that are rooted in EHR data. Our COMET model, which learned from over 809 million EHR features from almost 320,000 patients using pretraining, had superior performance metrics compared to models generated using EHR data alone, proteomics data alone, or both EHR and proteomics data combined without pretraining. This finding highlights the synergistic value of an integrative approach that utilizes multimodal data to generate biological insights into disease. Applying COMET may improve predictive modeling and generate novel biological insights without the need to increase the sample size of omics studies, which is attractive given that increasing sample size can be prohibitively expensive. COMET can be used as an initial feature selection step to identify proteins exhibiting nonlinear or dynamic expression patterns that traditional differential expression bioinformatic tools may not readily capture, as these tools are optimized to detect linear or monotonic changes (**Figure 5**; top panel). By subsequently validating these multimodal AI-prioritized proteins in an independent validation cohort (**Figure 5**; bottom panel), our workflow integrates the complementary strengths of both approaches. This two-stage strategy narrows the protein search space in a biologically informed manner and enhances the detection of robust candidates that might otherwise be overlooked in conventional omics analyses relying solely on univariate differential expression methods.

**Figure 5.**
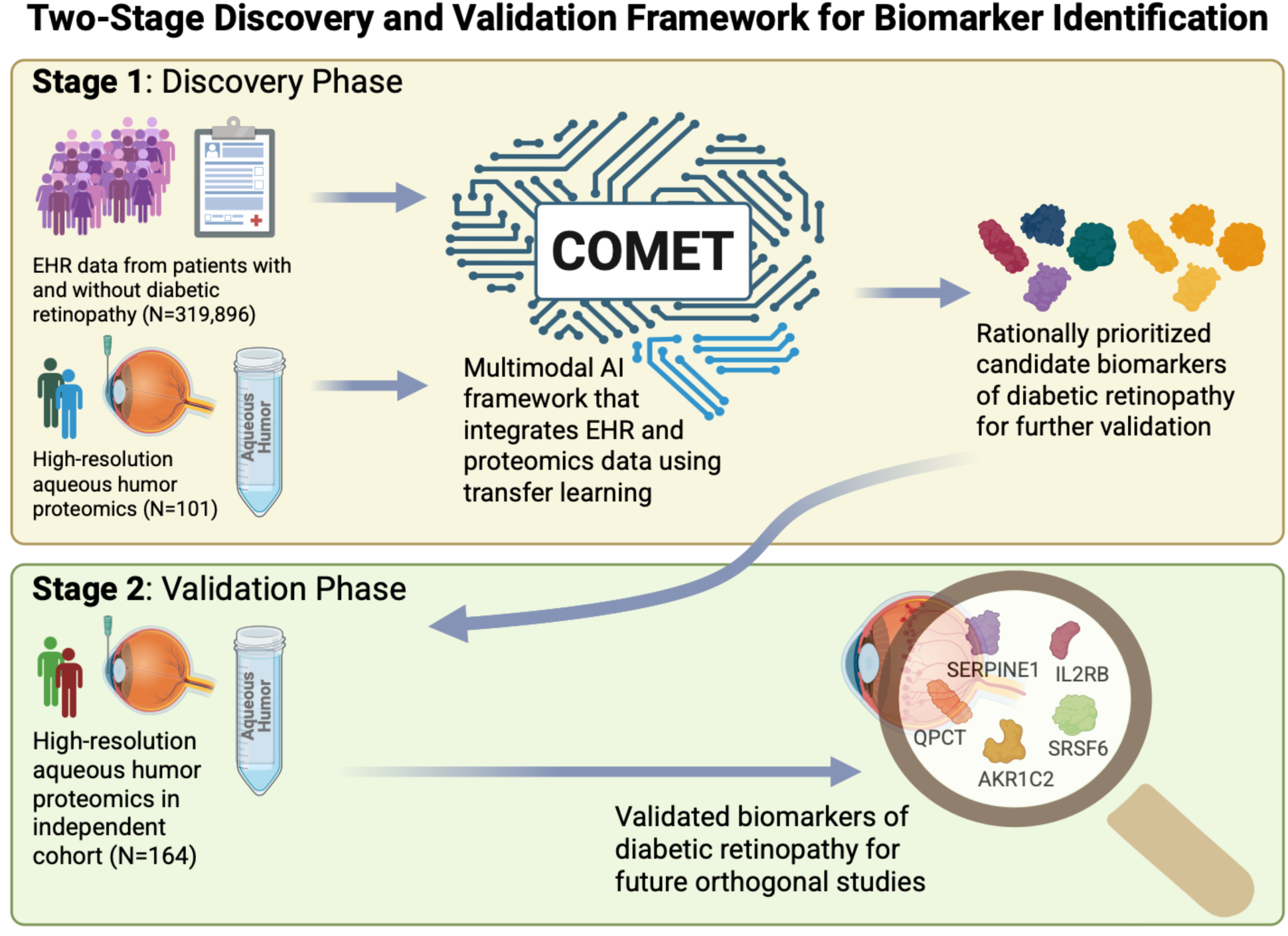
Schematic depicts our proposed two-stage framework for biomarker discovery. In the first stage (top panel), multimodal AI is used to integrate electronic health record (EHR) data from large cohorts and high-resolution proteomics from a subset of this larger cohort to rationally prioritize candidate biomarkers of disease. Subsequently, candidate biomarkers are validated in a second stage (bottom panel) in an independent cohort to identify robust, clinically relevant protein biomarkers of disease that can be investigated further in orthogonal, mechanistic studies.

Another potential advantage of integrating proteomics data with EHR data is the ability to capture the underlying disease heterogeneity in an agnostic manner and to enable the identification of protein-disease feature associations that may pave the foundation for future novel therapeutic strategies. For example, in the present study, correlational analysis in our cohort revealed associations between NOTCH2 and DME; NOTCH signaling has been suggested to have a possible pathologic role in vascular permeability and, thereby, in contributing to DME ^22^. Remarkably, in our study, this association was identified without any a priori hypotheses and without requiring any prespecification in study recruitment or design. We speculate that integrated EHR-proteomic approaches may enable us to explore these types of disease feature-protein associations in an unbiased fashion to enhance biological discovery. Additionally, this integrated approach may aid in identifying previously unrecognized systemic-ocular disease associations that may aid in therapeutics development.

Post hoc analysis of the pretrained model prioritized several proteins that had the higher relative feature importance in the pretrained model compared to the baseline model. Since they are more closely linked to the EHR data, these proteins may have more biological relevance and are, thus, more likely to be drivers of DR pathobiology. Supporting this hypothesis, most of these proteins have been identified in prior studies as having a possible role in DR pathobiology but have not been the focus of therapeutic design and testing to our knowledge. For example, elevated systemic levels of SERPINE1 are a hallmark of insulin-resistance syndrome and have been associated with obesity, insulin resistance, and increased vascular risk ^35,36^. Mice lacking SERPINE1 exhibit a mild hyperfibrinolytic phenotype ^37^. In mouse models of pathologic retinal angiogenesis, SERPINE1 promotes abnormal angiogenesis ^38^. In patients with diabetes, SERPINE1 is implicated in retinal microvascular disease, a key feature of DR ^39^. Moreover, elevated levels of SERPINE1 have been detected in vitreous biopsies and neovascular tissue from patients with PDR, suggesting that SERPINE1 may represent a novel therapeutic for modulating pathological angiogenesis ^40^.

While no studies to date have directly linked QPCT to DR, it has been investigated as a potential therapeutic target for neurodegenerative and inflammatory diseases ^41^. In Alzheimer’s disease (AD), QPCT contributes to the formation of toxic amyloid beta variants ^42^.

Varoglutamstat, a small-molecule QPCT inhibitor, has been studied both as a monotherapy and in combination with anti-amyloid beta antibodies ^43,44^. These findings suggest that QPCT may have previously unrecognized relevance in DR and could represent a novel biomarker for further investigation.

AKR1C2 has been reported to be differentially expressed in retinal fibrovascular membranes from patients with PDR ^45^, suggesting a possible role of neurosteroid metabolism in DR pathogenesis. In other ocular contexts, AKR1C2 expression is elevated in optic nerve head astrocytes from patients with glaucoma ^46^. In vitro studies have shown that AKR1C expression increases under elevated hydrostatic pressure in optic nerve head astrocytes, potentially leading to enhanced neurosteroid production and glaucomatous neurodegeneration ^46,47^.

Conversely, in models of dry eye disease, AKR1C2 has a protective role by mitigating oxidative stress-medicated ferroptosis ^48^. These findings suggest that AKR1C2 may have context-dependent functions in ophthalmic disease and could represent a novel target for further investigation in DR.

In the context of diabetes, retinal IL2 levels have been found to be significantly elevated in murine models ^49^. A recent meta-analysis of gene expression datasets reported significantly lower IL2RB expression in retinal tissues from patients with non-proliferative DR (NPDR) compared to fibrovascular membranes from patients with PDR ^50^. Beyond its immunological role, IL2 is also a key neuroregulatory cytokine ^51^ and has demonstrated neuroprotective effects in experimental models ^52,53^. Together, these findings suggest that IL2 signaling may contribute to the immunologic and neurodegenerative mechanisms underlying DR progression.

SRSF6 has been extensively studied in solid and hematopoietic malignancies ^54–56^. To date, no studies have specifically examined the role of SFSF6 in the eye. However, in other diseases, SFSF6 has been implicated in the post-transcriptional regulation of vascular endothelial growth factor (VEGF) ^57,58^, a molecule central to the pathogenesis of DR ^59^. These findings suggest that SFSF6 may contribute to DR through the modulation of VEGF expression, representing a potential unexplored molecular mechanism.

Using multimodal AI and EHR data improves our ability to translate omics data and may overcome previous challenges of using large omics studies to enhance care in the era of precision medicine ^60^. Other groups, for example, have harnessed the power of multimodal omics and AI to discover systemic disease associations with retinal thickness measured with optical coherence tomography ^61^. Further studies integrating omics and EHR data may allow for the prioritization of the most clinically relevant proteins and may provide concrete, biological insight into associations identified in larger retrospective cohort studies performed using claims data, such as the association between coronary heart disease and DR ^62^. Cumulatively, our findings highlight the synergistic value added by integrating EHR and omics data.

Another important finding is that the COMET framework improves modeling of EHR data by making it more grounded in the underlying biology. In the baseline model, there were no significant associations between proteins and the EHR latent representations; however, pretraining the model using the COMET pipeline led to an improved EHR latent representation that had over 4,000 significant pairs of associations between proteins and EHR latent representations. Incorporating proteomics data, even from a relatively small subset of the entire study population, teaches the model a more biologically meaningful EHR latent representation.

The final interesting finding is that when using TEMPO ^18^ to trace the identified proteins back to their cellular origin within the eye, we noted that the proteins originated from diverse ocular cell types, including not only retinal neurons but also immune and vascular cells. This finding highlights the complex pathobiology underlying DR and supports that there is likely a cell-autonomous component of disease that goes beyond systemic burden of disease. This may explain, in part, why there can be discordance between the systemic versus organ-specific burden of disease.

We acknowledge the limitations of our study. In this study, we analyzed relatively few patients with DR and, as such, run the risk of generating an overfitted model. In addition, DR is a heterogeneous disease with many different stages. Our study attempts to control for this since DR stage is included as an EHR feature within the analysis, but it remains possible that current DR staging is imprecise and, as such, our results could be biased by our specific study population. Nonetheless, our findings serve as a foundation for future larger studies that may allow us to perform similar analyses across different disease stages that may provide novel biological insights into the drivers of different stages of DR. One other important avenue for validation would be to perform similar studies at other institutions that have large-scale EHR databases available to test the generalizability of this multimodal AI approach. Additionally, there are known batch effects that are an inherent limitation of proteomics studies. Though we did not observe any batch effects within the present SomaLogic dataset, it is essential for these candidate protein biomarkers to be validated in future orthogonal studies utilizing in vitro and animal models to confirm their mechanistic role in DR.

In conclusion, transfer learning enables us to fuse EHR and proteomics data to generate novel biological insights in DR. We demonstrate that integration of EHR and proteomics data enhances biological discovery since it can identify novel disease feature-protein associations, highlight the most disease-relevant proteins of interest, and improve modeling of EHR data. As EHR data becomes more readily available and accessible and multimodal omics data become commonplace, integrative approaches will be necessary to make full use of the data. These innovative approaches are necessary to fully elucidate the pathobiology of blinding diseases and, one day, lead to novel therapeutic approaches.

## METHODS

### Subjects

The study protocol was approved by the Institutional Review Board (IRB) of the Stanford Research Compliance Office, was compliant with Health Insurance Portability and Accountability Act (HIPAA) policies, and adhered to the Declaration of Helsinki. All subjects underwent thorough discussion of expectations, risks, benefits, and alternatives, and provided informed consent prior to being included in the study. Aqueous humor biopsies were obtained at the time of planned ocular surgery. In brief, approximately 100 microliters of undiluted aqueous humor were manually aspirated from an anterior chamber paracentesis at the time of planned ocular surgery and was immediately frozen on dry ice and stored at –80°C prior to analysis.

There were two separate cohorts in this study: an initial discovery cohort (N=101) and a subsequent validation cohort (N=164). A case-to-control ratio of approximately 1:6 for the discovery cohort was selected to maximize statistical power in this large-scale association study while balancing costs ^63,64^. A similar ratio of relatively more cases compared to controls was also used for the validation cohort. The proteomics data from the discovery cohort (N=101) was previously used in other analyses that are distinct from that performed in the present study ^18^.

Though the patients in the validation cohort were also treated at Stanford and, therefore, likely part of the STARR OMOP database, we were not able to identify their EHR data without special permission since the database information is de-identified.

### Proteomic Profiling of Liquid Biopsies

We performed proteomic profiling as described previously ^18^. In brief, aqueous humor samples were analyzed with an aptamer-based proteomics assay (SomaScanⓇ Assay v4.1) and processed as described previously.

### Extracting and Processing EHR Data

We obtained EHR data from the **STA**nford **R**esearch data **R**epository (STARR) **O**bservational **M**edical **O**utcomes **P**artnership (OMOP) database ^65^, extracting patient information from five key tables: measurements, observations, drug exposures, condition occurrence, and procedure occurrence. For patients in the DR proteomics cohort, we analyzed their complete medical history up until the date of their aqueous humor liquid biopsy. For the other patients, we used data up until their time of eye disease onset as defined by first occurrence of any of the following condition concept identifiers: 45770830, 43530685, 45757435, 45763584, 45770881, 43530656, 376683, 380096, 4227210, 4266637, 37016179, 4174977, 36674200, 36674199, 35626042, 35626039, 35626038, 4290822, 377552, or 374338.

To transform raw EHR data into meaningful features for our deep learning model, we developed a sophisticated embedding approach as previously described ^17^. First, we gathered all unique concept codes across our EHR tables. We then organized these codes chronologically by patient and day, treating each day’s medical events as a collection of related concepts. Since precise timestamps within a day are often unreliable for medical records, we randomized the order of same-day events. Using word2vec, we created rich 400-dimensional embeddings for each medical concept. We built separate embedding models for our pretraining and proteomics cohorts to capture their unique characteristics. To summarize each patient’s daily medical events, we averaged the embeddings of all codes recorded that day. This process generated a series of daily summary vectors that capture the complexity of each patient’s medical journey. Our model processes the most recent 32 days of these summaries.

### COMET Architecture

COMET has been described in detail previously ^17^. In brief, our deep learning framework adapts to both regression and classification tasks while maintaining three core components: EHR analysis, omics processing, and integrated prediction. The EHR component employs a recurrent neural network (RNN) with gated recurrent units to analyze temporal patterns, followed by a linear transformation layer that generates EHR-based predictions. We optimized the network’s structure through hyperparameter tuning of layer count, hidden dimensions, and dropout rates. The omics component transforms molecular data through a single linear layer, while a separate joint layer combines EHR patterns with omics data to capture interactions between these different data types. The final prediction emerges from a bias-free linear layer that weighs and combines predictions from all three components, followed by a sigmoid, with the network optimized using binary cross entropy loss. We have previously developed a transformer-based variant of this architecture (COMET Transformer), which is another time-aware deep learning architecture. COMET Transformer was found to yield similar results, though slightly underperforming the RNN-based architecture.

### COMET Cross-Validation

To determine the COMET hyperparameters, we use a threefold cross-validation and grid search. Within the two training folds, we take 20% of the data as a test set for early stopping with a patience of 5 epochs. We assess the performance of each hyperparameter set via grid search on the validation set and choose the hyperparameter set that gives the lowest average loss on the validation sets across the threefold cross-validation.

These hyperparameters are used for all the subsequent experiments, including those with different train, test, and validation splits. For the EHR part of the network, the parameter grid is as follows: learning_rate, {1 × 10–1, 1 × 10–2, 1 × 10–3, 1 × 10–4}; dropout, {0.1, 0.2, 0.3, 0.4, 0.5}; lr_decay, {1 × 10–1, 1 × 10–2, 1 × 10–3, 1 × 10–4}; layers, {2, 4}; hidden_dim, {400, 800}. The batch size is fixed at 512 for the pretraining cohort and 16 for the omics cohort. For the proteomics-only experiment, we separately optimize the learning rate and lr_decay from the same range. From the joint experiments, we optimize learning rate, dropout and lr_decay, but fix the number of layers and hidden dimension as the optimal weights chosen from the EHR-only experiments. When we transfer the weights, we fix the number of layers and dropout as the optimal values from the pretraining experiments but still optimize learning rate and lr_decay from the aforementioned range. Using these hyperparameters, we performed twenty-five iterations of each experiment using different train, test, and validation splits. The training set is 70% of the data, and the test and validation sets are 15% each. We again use the test set for early stopping with a patience of 5 epochs. To compute the final performance metrics, we averaged the predictions from the validation set across all the twenty-five iterations and used those averaged predictions to compute a final prediction for each patient. We calculated 95% confidence intervals (CI) for AUROC and AUPRC by performing one thousand iterations of bootstrapping.

### Post Hoc Statistical Analysis

We performed post hoc data visualization and statistical analysis using R version 4.4.1 and RStudio version 2024.04.2+764. To compare means between two groups, we performed an unpaired t-test after verifying that the data followed a normal distribution by visual inspection. The differential expression analysis focused on the fifteen proteins identified by the COMET model. Differential expression analysis was carried out using the *limma* package, incorporating robust linear modeling. To account for potential batch variability, we used external calibrators to scale the signal across different experimental batches. Within each sample type, data were median-normalized to ensure comparability. To assess the presence of residual batch effects, we performed principal component analysis (PCA), generated unsupervised hierarchical heatmaps, and examined boxplots of normalized intensities. These analyses did not reveal any obvious batch effects within the present dataset. P values were corrected for multiple comparisons using the Benjamini-Hochberg procedure, and proteins with an adjusted p-value < 0.05 were considered statistically significant.

### Power Calculations

We calculated the post hoc power of the discovery cohort by using the median log fold change of 2.6 (d=1.4) among the fourteen of the 15 proteins highlighted by COMET that were statistically significant and the empirical standard deviation of 1.9. Our post hoc power was 99.7%. We used the magnitude of effect size from the discovery cohort to estimate the sample size needed for the validation cohort. To achieve 90% power with this estimated effect size of d=1.4, we calculated that we would require 127 controls and 7 patients with DR. To ensure rigor, the validation cohort exceeded this sample size and consisted of N=164 participants (17 with DR and 147 without).

## Data Availability

The proteomics data is available upon request to the corresponding author. The EHR data and proteomics data from patients cannot be shared publicly due to Stanford University policies.

## Code Availability

The code is available at https://github.com/samson920/COMET_DR/.

## Material Availability

This study did not generate new unique reagents.

## Data Availability

The proteomics data is available upon request to the corresponding author. The code is available at https://github.com/samson920/COMET_DR/.

## Acknowledgements

We thank Aarushi Kumar and Artis Mongague, MD, PhD, for assistance with sample collection and MaryAnn Mahajan for editorial assistance. VBM is supported by NIH grants (R01EY037830, R01EY031360, R01EY030151, and P30EY026877), the Alan Adler Center for Ophthalmic Oncology, and Research to Prevent Blindness. JBL was supported by the VitreoRetinal Surgery Foundation and the Heed Ophthalmic Foundation. SJM was supported by the NSF GRFP (DGE-1656518). GV is supported by NIH grant R38EY037090. This research used data or services provided by STARR, “STAnford medicine Research data Repository,” a clinical data warehouse containing live Epic data from Stanford Health Care, the Stanford Children’s Hospital, the University Healthcare Alliance and Packard Children’s Health Alliance clinics, and other auxiliary data from Hospital applications, such as Radiology PACS. The STARR platform is developed and operated by the Stanford Medicine Research Technology team and is made possible by the Stanford School of Medicine Research Office. The funding organizations had no role in design and conduct of the study; collection, management, analysis, and interpretation of the data; preparation, review, or approval of the manuscript; and decision to submit the manuscript for publication.

## Author Contributions

VBM and NA had full access to all the data in the study and take responsibility for the integrity of the data and the accuracy of the data analysis. Study concept and design: SJM, NA, VBM. Acquisition of data: SJM, VBM, NA. Analysis and interpretation of data: SJM, JBL, MC, GV, PM, NA, VBM. Drafting of the manuscript: JBL, SJM, VBM. Critical revision of the manuscript for important intellectual content: all authors. Statistical analysis: SJM, JBL, NA. Obtained funding: VBM, NM. Administrative, technical, and material support: NA, VBM, PM. Study supervision: NA, VBM.

