## Supplementary Data 2 for "Multimodal AI fuses proteomic and EHR data for rational prioritization of protein biomarkers in diabetic retinopathy"

**Supplemental Table 2**. Demographic and clinical information of participants in the validation cohort.

| **Characteristic** | **Control (N=147)** | **Diabetic Retinopathy (N=17)** |
| --- | --- | --- |
| Age, mean (SD) | 68.8 (12.1) | 62.2 (10.4) |
| Sex |  |  |
| Male, n (%) | 58 (39.5%) | 7 (41.2%) |
| Female, n (%) | 89 (60.5%) | 10 (58.8%) |
| Diabetic Retinopathy |  |  |
| None, n (%) | 147 (100%) | 0 (0%) |
| Non-Proliferative, n (%) | 0 (0%) | 7 (41.2%) |
| Proliferative, n (%) | 0 (%) | 10 (58.8%) |
